# Effect of Pathway-specific Polygenic Risk Scores for Alzheimer’s Disease (AD) on Rate of Change in Cognitive Function and AD-related Biomarkers among Asymptomatic Individuals

**DOI:** 10.1101/2023.01.30.23285142

**Authors:** Yuexuan Xu, Eva Vasiljevic, Yuetiva K. Deming, Erin M. Jonaitis, Rebecca L. Koscik, Carol A. Van Hulle, Qiongshi Lu, Margherita Carboni, Gwendlyn Kollmorgen, Norbert Wild, Cynthia M. Carlsson, Sterling C. Johnson, Henrik Zetterberg, Kaj Blennow, Corinne D. Engelman

## Abstract

**Background:** Genetic scores for late-onset Alzheimer’s disease (LOAD) have been associated with preclinical cognitive decline and biomarker variations. Compared with an overall polygenic risk score (PRS), a pathway-specific PRS (p-PRS) may be more appropriate in predicting a specific biomarker or cognitive component underlying LOAD pathology earlier in the lifespan.

**Objective:** In this study, we leveraged 10 years of longitudinal data from initially cognitively unimpaired individuals in the Wisconsin Registry for Alzheimer’s Prevention and explored changing patterns in cognition and biomarkers at various age points along six biological pathways.

**Methods:** PRS and p-PRSs with and without apolipoprotein E (*APOE*) were constructed separately based on the significant SNPs associated with LOAD in a recent genome-wide association study meta-analysis and compared to *APOE* alone. We used a linear mixed-effects model to assess the association between PRS/p-PRSs and global/domain-specific cognitive trajectories among 1,175 individuals. We also applied the model to the outcomes of cerebrospinal fluid biomarkers for beta-amyloid 42 (Aβ42), Aβ42/40 ratio, total tau, and phosphorylated tau in a subset. Replication analyses were performed in an independent sample.

**Results:** We found p-PRSs and the overall PRS can predict preclinical changes in cognition and biomarkers. The effects of p-PRSs/PRS on rate of change in cognition, beta-amyloid, and tau outcomes are dependent on age and appear earlier in the lifespan when *APOE* is included in these risk scores compared to when *APOE* is excluded.

**Conclusion:** In addition to *APOE*, the p-PRSs can predict age-dependent changes in beta-amyloid, tau, and cognition. Once validated, they could be used to identify individuals with an elevated genetic risk of accumulating beta-amyloid and tau, long before the onset of clinical symptoms.

## Introduction

Late-onset Alzheimer’s disease (LOAD) is an age-related neurodegenerative disease that is clinically manifested by a progressive deterioration of cognitive function, memory, and social ability. Abnormal accumulation of proteins such as β-amyloid (Aβ) and tau are two hallmarks that play important roles in LOAD pathology long before the clinical symptoms of neurodegeneration are evident[1]. Under the amyloid hypothesis, an imbalance between Aβ clearance and Aβ production is considered the underlying cause for the initiation of LOAD through the formation of extracellular senile plaques in the brain [2]. Previous studies have provided evidence that neurobiological pathways, such as β-amyloid precursor protein (APP) processing, altered cholesterol metabolism, endocytosis, and tau pathology, are closely linked to Aβ production and clearance [3–9]. Tau, on the other hand, is hypothesized to trigger the progression of LOAD by forming insoluble filaments and accumulating intracellular neurofibrillary tangles of hyperphosphorylated tau in the brain. These accumulations block axonal transport and finally harm the synaptic communications between neurons. In addition to the four pathways affecting Aβ, neurobiological pathways of LOAD that are related to tau accumulation among LOAD patients include immune response and axonal development [3–7,9–12].

Genetics play a major role in LOAD. LOAD is highly polygenic, and the heritability estimates from twin studies range from 58% to 79% [13]. The *apolipoprotein E* (*APOE*) gene is the strongest known genetic risk factor for LOAD, with the *APOE* ε4 allele conferring increased risk and the *APOE* ε2 allele conferring a protective effect relative to the *APOE* ε3 allele[14]. A meta-analysis of genome-wide association studies (GWAS), which included more than 94,000 individuals with European genetic ancestry, confirmed 20 previously reported risk loci and discovered five novel, susceptibility single-nucleotide polymorphisms (SNPs) [15]. However, except for *APOE*, most of the discovered genetic variants only exhibit tiny effects on the risk of LOAD, and therefore the prediction from any single genetic variant is limited. Polygenic risk scores (PRSs), on the other hand, sum the effects of multiple independent SNPs and convert the overall genetic burden to a single score. This score has been found to serve as a good predictor of disease risk [16–18]. Although an overall PRS that combines genetic variants across the genome is more commonly used and may be more powerful in the prediction of the overall cognitive status or LOAD risk, a pathway-specific polygenic risk score (p-PRS) that sums individual SNPs under a specific neurobiological pathway may be more appropriate in predicting a specific biomarker or cognitive component (such as the beta-amyloid 42/40 ratio, phosphorylated tau, or executive function) underlying LOAD etiology[19,20].

To date, a constellation of studies has been published to examine the prediction performance of p-PRS on LOAD disease risk, cognitive decline, and biomarker variation among people with or without LOAD; however, the study findings are mixed. Previous research from our group examined the prediction performance of p-PRSs under three pathways on cognition, Aβ burden in the brain as measured with Pittsburgh compound B Positron Emission Tomography (PiB-PET), and cerebrospinal fluid (CSF) Aβ and tau using a prospective cohort of 1,200 asymptomatic individuals[19]. We found that p-PRSs under all three pathways were not predictive of the global or domain-specific cognitive scores, whereas p-PRSs in the Aβ and cholesterol pathways were good predictors of variations of PiB amyloid accumulation and CSF Aβ and tau. However, the predictive performance was significantly reduced with the exclusion of the *APOE* variants. Another study investigated the effect of p-PRSs under seven pathways on cortical thickness using a longitudinal population cohort of 544 individuals [21]. Significant associations with cortical thickness were discovered in the APP metabolism, cholesterol metabolism, and endocytosis pathways when *APOE* was included; however, only the APP metabolism pathway remained significant after adjustment for the *APOE* variants. A recent study estimated the risk of LOAD among 1,779 Dutch individuals using p-PRSs in five major pathways involved in LOAD [22]. They found that all p-PRSs except for angiogenesis were significantly associated with increased risk of LOAD, regardless of adjustment for the *APOE* variants. Several reasons may exist for the discrepant results among the existing LOAD-related p-PRS analyses, but it is likely because different LOAD outcomes are used and the methods for pathway-gene-variant mapping did not draw from a comprehensive body of literature. In addition, age is the strongest factor associated with variation in the endophenotypes and cognitive decline, but it was not considered as more than a covariate in the existing literature when assessing the predictive performance of p-PRSs on cognition and LOAD-related biomarkers. A recent study leveraging a 25-year longitudinal cohort of non-demented individuals showed that the overall LOAD genetic risk on cognitive decline is age-related during the life course [23].

In the present study, we updated findings from Darst et al. (2017)[19] with five additional years of follow up data from an ongoing longitudinal cohort of cognitively healthy adults enriched for a parental history of AD from the Wisconsin Registry for Alzheimer’s Prevention (WRAP) to explore the potential of p-PRSs in the prediction of cognitive decline and changes in LOAD-related biomarkers over time. Specifically, after a comprehensive review of the existing literature on the LOAD disease pathways and functions of the genes identified by the recent case-control GWAS meta-analysis, we constructed weighted p-PRSs for APP metabolism, cholesterol metabolism, endocytosis, tau pathology, immune response, and axonal development. For each p-PRS, we tested its association with a global cognitive composite score (Preclinical Alzheimer Cognitive Composite – 3 (PACC-3)), domain-specific cognitive composite scores (Immediate Learning, Delayed Recall, and Executive Function), and biomarkers of Aβ accumulation (CSF Aβ42 and CSF Aβ42/40 ratio), neurodegeneration (CSF total tau [T-tau)), and tau pathology (CSF phosphorylated tau [P-tau)), while taking heterogeneity in genetic risk by age into account. To check the robustness of the results, we further performed a replication analysis using an independent sample of cognitively healthy individuals from the Wisconsin Alzheimer’s Disease Research Center (ADRC).

## Methods

### Study participants

Data leveraged in this study originated from WRAP, an ongoing longitudinal prospective cohort study of middle-aged adults who were cognitively healthy at enrollment and spoke English (N > 1,500). WRAP is enriched for participants with a parental history of clinical AD, increasing the proportion of individuals who will experience AD pathology and cognitive decline during the course of the study. The details of the study design have been described elsewhere [24]. The WRAP study began recruiting participants in 2001 with an initial follow-up after four years and subsequent follow-up every two years. In general, the participants were between 40 and 65 years old at baseline. Siblings of WRAP participants were allowed to enroll. Participants were given an extensive battery of neuropsychological tests at each visit. The maximum number of WRAP visits available at the time of analysis, using freeze20May, was seven. In the present study, the sample was limited to self-reported non-Hispanic white (NHW) participants to match the race and ethnicity of the participants in the GWAS meta-analysis from which the weights for the PRS were drawn. We excluded data from the baseline WRAP visit because the cognitive outcome examined in this study cannot be computed using the neuropsychological tests administered in the first WRAP visit. Data from the seventh visit of WRAP were excluded because data collection in the seventh visit is ongoing and data from this visit were only available for less than 50 participants. Compared to the previous p-PRS study on the LOAD-related outcome using WRAP[19], the present study includes additional data from approximately two more WRAP visits per participant. This study was conducted with the approval of the University of Wisconsin Institutional Review Board, and all subjects provided signed informed consent before participation.

### Neuropsychometric assessments

Participants were given a battery of neuropsychological tests for the assessment of cognitive function at each WRAP visit. In the present study, we measured the overall cognitive performance using the PACC-3 score based on work by Donohue and colleagues[25]. Specifically, this WRAP composite score is computed by standardizing and averaging performance from three tests that assess the memory and executive function of participants: Rey Auditory Verbal Learning (RAVLT; Trials 1-5), Wechsler Memory Scale-Revised (WMS-R) Logical Memory II total score (LMII), and Wechsler Adult Intelligence Scale-Revised (WAIS-R) Digit Symbol Coding total items completed in 90 seconds [26]. In addition to the overall cognitive performance, we also examined domain-specific cognitive performance for immediate learning, delayed recall, and executive function [27]. The immediate learning domain-specific composite score was derived from RAVLT total trials 1-5, WMS-R Logical Memory I total score (LMI), and Brief Visuospatial Memory Test-Revised (BVMT-R) immediate recall score. A delayed recall domain-specific composite score is constructed based on the sum of the RAVLT long-delay free recall score, WMS-R logical memory delayed recall score, and BVMT-R delayed recall score. The executive function domain-specific composite score is obtained based on Trail-Making Test part B (TMT-B) total time to completion, Stroop Neuropsychological Screening Test color-word interference total items completed in 120 seconds, and WAIS-R Digit Symbol Coding. All three domain-specific cognitive composite scores are calculated by averaging standardized test scores as previously described[27]. The z-score for TMT-B was multiplied by -1 before the inclusion into the composite so that higher z-scores indicate better performance for all tests.

### CSF collections, quantification, and analysis

A subset of WRAP participants consented to a lumbar puncture to obtain CSF. Details and methods for the WRAP CSF processing have been described elsewhere [28]. In brief, 22 mL of CSF were collected through gentle extraction and combined into a 30 mL polypropylene tube. All CSF samples were processed in one batch at the Clinical Neurochemistry Laboratory at the Sahlgrenska Academy of the University of Gothenburg in Sweden using the Roche NeuroToolKit robust prototype assays (Roche Diagnostics International Ltd, Rotkreuz, Switzerland) under strict quality control procedures as previously described [29]. CSF measurements examined in the present study include Aβ42, Aβ42/40 ratio, T-tau, and P-tau. Previous studies have indicated that CSF Aβ42 levels are negatively associated with amyloid burden; however, higher levels of CSF T-tau and P-tau are signals of increased tau pathology [30]. We also examined the CSF Aβ42/40 ratio because it has greater predictive and diagnostic power in early diagnosis of LOAD compared to the individual biomarker CSF Aβ42 alone [31].

### DNA collection, genotyping, and quality control

Details about genomic data collection have been described elsewhere [19,32]. Briefly, we used the PUREGENE DNA Isolation Kit to extract DNA from whole blood samples, and then we used UV spectrophotometry to quantify DNA concentrations. Of the 23 SNPs included in the analysis, 21 were genotyped in 1,448 individuals using competitive allele-specific PCR-based KASP™ genotyping assays (LGC Genomics, Beverly, MA). Duplicate quality control (QC) samples had 99.9% genotype concordance, and all discordant genotypes were set to missing. The QC was carried out using PLINK v1.07[33]. Individuals with high missingness of alleles (>10%) were removed. A total of 1,415 individuals remained in the sample after QC procedures. All 21 SNPs had call rates >95% and were in Hardy-Weinberg equilibrium (HWE).

Two SNPs (rs12459419 from *CD33* and rs593742 from *ADAM10*) that were not genotyped by the KASP™ assays were extracted from genome-wide genotyping performed using the Illumina Infinium Expanded Multi-Ethnic Genotyping Array (MEGA^EX^) at the University of Wisconsin Biotechnology Center. Standard quality control procedures for genome-wide data were performed and have been described previously[32]. Genotypic data from individuals of European genetic ancestry were then imputed using the Michigan Imputation Server and the Haplotype Reference Consortium (HRC) reference panel. SNPs rs12459419 and rs593742 were imputed with a high quality (R^2^ > 0.8). A total of 1,198 individuals with data for all 23 SNPs remained after QC.

### Mapping variants to pathways

To address the limitations in the traditional pathway-gene-variant mapping method in p-PRS studies that did not draw from a comprehensive body of literature (often relying on a single paper) and relieve concerns about the validity (e.g., overweighting or underweighting a particular variant) of the novel approach in the p-PRS construction proposed recently [22], we combined the merits of these two approaches and designed a new but conservative strategy to map genetic variants to various LOAD pathways. First, we comprehensively browsed pathways explored in the past LOAD-related p-PRSs studies published in peer-reviewed journals between 2017 and early 2020 to determine pathways that had been widely explored [19,21,22]. After a review of the literature, we included six pathways in the present analysis: APP metabolism, cholesterol metabolism, endocytosis, tau pathology, immune response, and axonal development. Second, we narrowed our focus to the genes with variants that were genome-wide significant, as identified by the most recent and largest International Genomics of Alzheimer’s Project (IGAP) GWAS meta-analysis on diagnosed AD [15]. In addition, we included genome-wide significant variants from three genes (*MEF2C, NME8*, and *CD33*) identified in previous GWAS meta-analyses [34–36]. These were widely mentioned in previous AD review papers and were marginally significant in Kunkle et al. (2019). Third, we extensively browsed recent review papers on LOAD pathology published between 2017 and early 2020, with the number of citations set to higher than 5 [3–9]. Then we counted the number of times the genes identified in step 2 presented in any of the specific pathways in the papers we reviewed. A specific gene was finally counted toward one of the pathways identified in step 1 only if more than 50% of the reviewed literature showed evidence that this gene belongs to that particular LOAD pathway. We finally included 22 genes in the main analysis under six pathways: APP metabolism (*CLU, SORL1, ABCA7, PICALM, ADAM10, APOE*), cholesterol metabolism (*CLU, SORL1, ABCA7, APOE*), endocytosis (*SORL1, ABCA7, PICALM, BIN1, CD2AP, PTK2B, FERMT2, SLC24A4, APOE*), tau pathology (*BIN1, FERMT2, CASS4, APOE*), immune response (*CLU, ABCA7, CR1, INPP5D,HLA-DRB1, TREM2, EPHA1, MS4A6A, CD33, MEF2C*), and axonal development (*EPHA1, FERMT2, CASS4, SPI1, NME8*) (Supplemental Figure 1).

### Polygenic risk score and pathway-specific polygenic risk scores

The *APOE* risk score was coded according to the odds ratios (ORs) of ε2, ε3, and ε4 genotypes based on rs7412 and rs429358 in the meta-analysis of *APOE* genotype frequencies from AlzGene [37]. Specifically, we constructed an *APOE* score using the ε2/ε2 genotype as the reference (ε2/ε2, OR=1): OR(ε2/ε3) = 1.38, OR(ε3/ε3) = 2, OR(ε2/ε4) = 4.45, OR(ε3/ε4) = 6.78, OR(ε4/ε4) = 25.84 [19]. Then, we log-transformed and added the score to the corresponding PRS/p-PRS. For genes other than *APOE*, the most significant variant from each of the 21 genes identified by the IGAP GWAS meta-analysis was used in the construction of PRS and p-PRS. PRS and p-PRS were calculated using the formula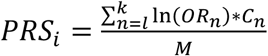, where *i* represents the *i*th individual whose PRS is calculated by summing all SNPs *n* in the pathway from the first SNP *l* to last SNP *k*; OR is the odds ratio of the risk allele for SNP *n* from the IGAP GWAS meta-analysis; C is the number of risk alleles for SNP *n* for individual *i*; and M represents the number of non-missing SNPs under each predetermined pathway observed in individual *i*. In addition to the p-PRS, an overall PRS by including all 22 genes was constructed to examine the overall genetic effect by summing SNPs in all pathways of LOAD being investigated on the outcome of interest. A higher PRS/p-PRS indicates a higher genetic risk for LOAD. Since the effect size of *APOE* alone is known to be large, we excluded *APOE* for the pathways that theoretically should include *APOE* to examine the p-PRS on the outcome beyond *APOE* alone. We also tested the association between the *APOE* score and the outcome of interest to quantify the effect of *APOE* alone. To facilitate comparison across various pathways, all PRS, p-PRSs, and *APOE* scores were standardized with a mean of 0 and a standard deviation of 1.

### Statistical analysis

We developed a set of linear mixed-effects models to examine the genetic association with cognitive outcomes and LOAD-related biomarkers while accounting for within-family (sibship) and within-individual correlations and missing data. All regression analyses related to linear mixed effects models were performed using the MIXED procedure implemented in Statistical Analysis Software (SAS) 9.4. Following the previous literature, we included random intercepts for family and study subjects [19,23]. WRAP investigators have reported the nonlinear effect of age on cognitive decline, and we therefore included a linear age, quadratic age, and cubic age in the model with cognitive outcomes to achieve better model fit [24]. For the biomarker analysis, we first used spaghetti plots to check the individual trajectory in the change of biomarkers by age and then determined the appropriate functional form of age based on the visualization of individual trajectories. To better model the dynamic relationship between aging, genetic risk, and LOAD-related outcomes, we further included an interaction term between PRS/p-PRS and all age terms to control for the potential age-related genetic risk on all outcomes of interest. In addition to the PRS/p-PRSs, age, and interaction between age and PRS/p-PRSs mentioned above, additional covariates include sex, education, practice effect (only adjusted in cognitive-related outcomes and quantified by the number of tests completed prior to the current test), and the first five genetic principal components[38]. We assessed the variance explained by each PRS/p-PRS and each interaction term between age and PRS/p-PRS using the incremental likelihood ratio 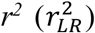 [39,40]. The significance of the interactions between each PRS/p-PRS and all age terms is evaluated by the likelihood ratio test (LRT). Upon discovering significant interactions, we further probe the nature of the interactions by investigating the conditional effect of each PRS/p-PRS at different age values and test them versus zero for significance using the simple slope approach. All simple slope estimates were calculated using the PLM procedure implemented in SAS 9.4.

### Replication analysis

We replicated all analyses performed in the WRAP sample within the Wisconsin ADRC, which began enrolling participants in 2009. Because the Wisconsin ADRC administered a different battery of neuropsychological tests compared to WRAP, we could only replicate our analyses for the overall cognitive performance (PACC-3) and CSF-related outcomes. We replicated our findings using two samples from the Wisconsin ADRC. The first sample is the IMPACT cohort, for which the enrollment criteria (age range of 45-65 at baseline, cognitively unimpaired, and enriched for a parental history of AD) are the most similar and comparable to the WRAP sample. To broaden the age range represented, we supplemented the IMPACT sample with the Wisconsin ADRC healthy older controls (HOCs), which includes people who are older than 65 at enrollment and do not meet the National Institute on Aging and Alzheimer’s Association criteria for mild cognitive impairment (MCI) or the National Institute of Neurological and Communicative Disorders and Stroke and Alzheimer’s Disease and Related Disorders Association (NINCDS-ADRDA) criteria for probable AD. We called this combined sample the All Healthy Controls (AHC) sample. All replication analyses in the Wisconsin ADRC were restricted to people who self-identified as NHW.

The Wisconsin ADRC administered a different battery of neuropsychological tests than WRAP, which resulted in a substantial missingness in the score of Logical Memory II Delayed Recall and Wechsler Adult Intelligence Scale-Revised, Digit Symbol. To make the best use of the current information, we consulted neuropsychologists in the Wisconsin ADRC and created a PACC-3-TMT score to replace PACC-3 in the replication analysis. Specifically, we converted the Craft Story score to an estimated Logical Memory score based on a published crosswalk table and followed the previous practice of substituting the Digit Symbol score with the total time to completion in the TMT-B test [26,41]. Since the published crosswalk table is only available for converting the Craft Story score to the Logical Memory score for the first five visits, we restricted our replication analysis using only data from the first five Wisconsin ADRC visits. The final PACC-3-TMT score was computed by standardizing and averaging the results from RAVLT, the estimated Logical Memory score, and the reversed coded TMT-B test results. The substantial missingness in the score of the Logical Memory score and Digit symbol makes it difficult to assess the correlation between PACC-3 and PACC-3-TMT in the Wisconsin ADRC; therefore, we used the same method to construct a PACC-3-TMT score in the WRAP and assessed the correlation between PACC-3 and PACC-3-TMT in the WRAP sample.

The Wisconsin ADRC employed the same methods of collection, processing, and quantification for the CSF data as those in WRAP [19]. Details about genomic data collection and QC have been described elsewhere [19,42]. Briefly, the top significant SNPs from 21 genes except for *ADAM10* were genotyped by LGC Genomics (Beverly, MA) using the same competitive allele-specific PCR-based KASP™ genotyping assays as in WRAP. All SNPs had call rates >95% and were in Hardy-Weinberg equilibrium (HWE). Two individuals with high missingness of alleles (>10%) were excluded from subsequent analyses. Only the APP metabolism pathway-specific PRS and overall PRS were affected by the exclusion of the *ADAM10* gene, but we expect the impact will be small due to the small effect size (β=-0.065) of the top significant SNP from *ADAM10*. The methods of constructing the PRS, p-PRS, and *APOE* scores are the same as those in WRAP analysis.

We utilized a linear mixed-effects model to examine the genetic association with the overall cognitive performance and LOAD-related biomarkers by accounting for within-individual correlations and allowing for missing data. All analyses were performed using SAS 9.4. All other statistical methods in the replication analyses are the same as those described in the WRAP analysis, except for the exclusion of genetic principal components as covariates because genome-wide data and, thus, genetic principal components for the full sample are not available in the Wisconsin ADRC genomic dataset.

## Results

### Descriptive statistics for samples and participants

Table 1 presents the demographic features for WRAP participants included in this study. Briefly, a total of 1,175 individuals with available genetic, cognitive, and demographic data remained in the sample for up to five visits (∼8 years) after the implementation of the inclusion criteria. A subset of 197 WRAP participants had CSF data for up to five visits. Demographic characteristics are comparable between the cognitive and CSF samples. The sample with CSF is slightly older at baseline than the full WRAP sample because WRAP CSF collection began later during the WRAP study. WRAP participants are generally highly educated, female, and enrolled at middle age, and a majority have a parental history of AD. The *APOE* score is not available for five participants in the full sample and one participant in the CSF sample because of missing allele information for either rs7412 and/or rs429358. Following the previous literature, we decided to keep these individuals in the analysis because data are available on other genetic variants that we are interested in, and the magnitude of missingness is small.

**Table 1.** Participant characteristics for WRAP at baseline

| Variable | Full sample (N=1,175) | Biomarker sample (N=197) |
| --- | --- | --- |
| Age | 58.57 (6.52) | 61.98 (6.64) |
| Education (years) | 15.81 (2.24) | 16.16 (2.14) |
| Gender (male) | 353 (30%) | 69 (35%) |
| Family history of AD | 858 (73%) | 142 (72%) |
| **Max visits** |  |  |
| 1 | 41 (4%) | 70 (36%) |
| 2 | 85 (7%) | 42 (21%) |
| 3 | 182 (15%) | 60 (30%) |
| 4 | 369 (31%) | 23 (12%) |
| 5 | 498 (42%) | 2 (1%) |
| **APOE genotypes** |  |  |
| $\epsilon 2/\epsilon 2$ | 4 (0.3%) | 0 (0) |
| $\epsilon 2/\epsilon 3$ | 96 (8%) | 21 (11%) |
| $\epsilon 3/\epsilon 3$ | 619 (53%) | 107 (54%) |
| $\epsilon 2/\epsilon 4$ | 39 (3%) | 6 (3%) |
| $\epsilon 3/\epsilon 4$ | 369 (31%) | 56 (28%) |
| $\epsilon 4/\epsilon 4$ | 43 (4%) | 6 (3%) |
| CSF Aβ42 (pg/mL) | N/A | 899.31 (391.24) |
| CSF Aβ42/40 | N/A | 0.06 (0.02) |
| CSF T-tau (pg/mL) | N/A | 208.82 (69.89) |
| CSF P-tau (pg/mL) | N/A | 18.34 (6.71) |

Mean (SD); n (%); Aβ : Beta-amyloid; P-tau: phosphorylated Tau; T-tau: total Tau.
“Max visits” refers to number of cognitive assessments of outcomes included in these analyses (left column) or lumbar puncture draws per participant (right column).

### Cognitive outcomes

Statistically significant interactions (*p* < 0.05) between the PRSs of all pathways and polynomial age were observed for all cognitive outcomes when *APOE* was included in the PRS (Figure 1a; regression output for first model in supplementary excel table; full regression outputs are available upon request). However, when *APOE* was excluded from the PRS, significant p-PRSs*polynomial age interactions were only observed under the endocytosis pathway for the immediate learning composite score; under APP metabolism, endocytosis, and tau pathology pathways for the delayed recall composite score; under endocytosis and tau pathology pathways for the executive function composite score; and under the endocytosis pathway for the PACC-3 composite score. Significant interactions between the overall PRS and polynomial age were observed for all cognitive outcomes when *APOE* was included in the PRS but were only observed for executive function and PACC-3 when *APOE* was excluded from the PRS.

**Figure 1.**
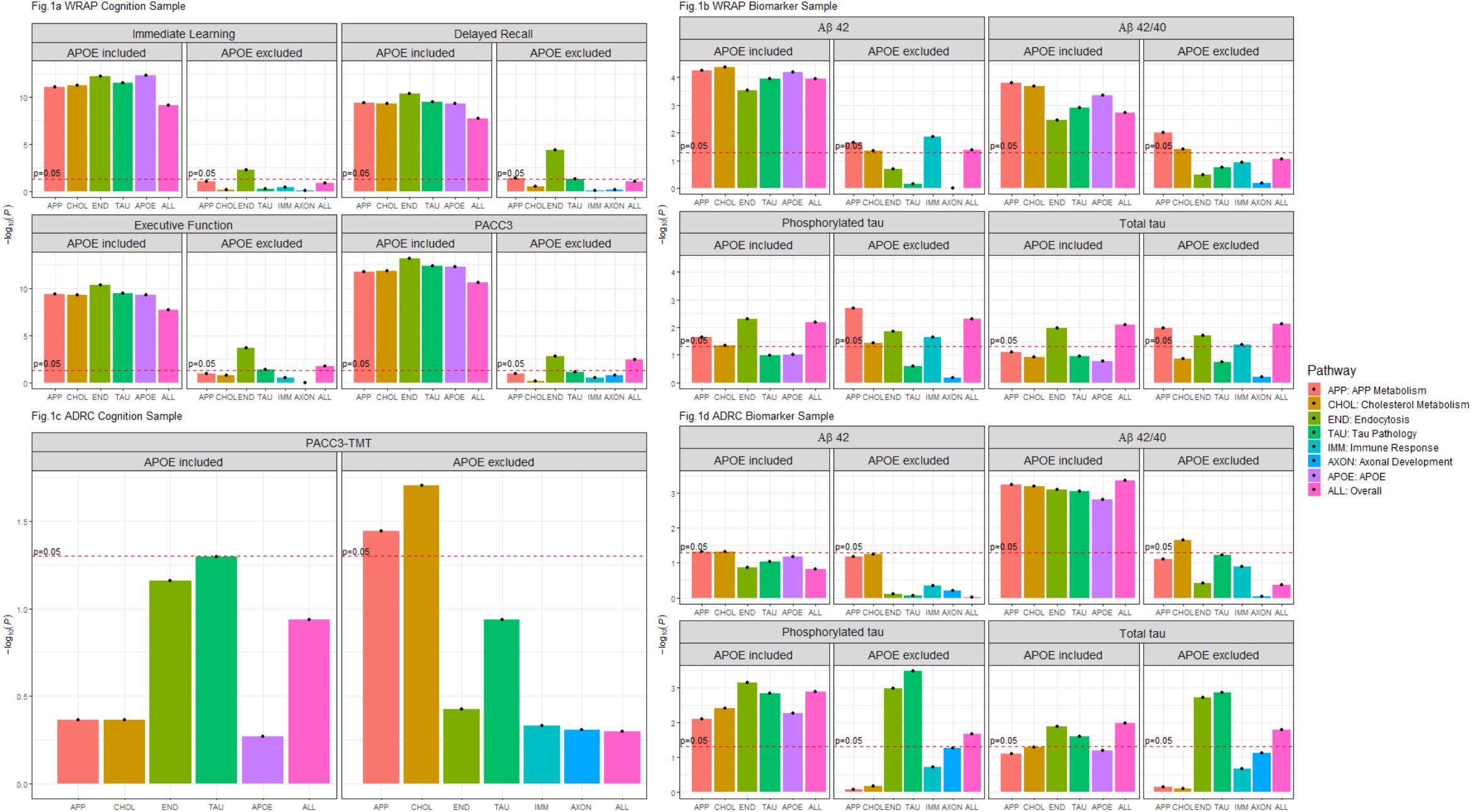
Likelihood ratio test results of the interactions between PRS/p-PRSs and polynomial age. presents the -log10(*P*) from the likelihood ratio tests for the interaction between polynomial age and genetic risk for all outcomes in WRAP and Wisconsin ADRC. The likelihood ratio test statistic is calculated as the ratio between the log-likelihood of the nested model (model without interaction terms) to the full model (model with polynomial age*genetic risk terms). All association analyses are performed using the linear mixed effect model and adjusted for within-individual and within-family correlation. In addition to p-PRSs/PRS, age (linear, quadratic, and cubic), and their interactions, additional covariates include gender, education years, practice effect, and the first five genetic principal components of ancestry.

Figure 2 presents the simple slope estimates of annual cognitive change by a one standard deviation change in genetic risk of PRS/p-PRS and by age in five-year increments between 50 and 80 years old for WRAP participants. In models in which *APOE* variants are included in the PRS, on average, the simple slope estimates of annual cognitive change are not statistically different from zero prior to age 65 for all pathways and every cognitive outcome. However, the simple slope estimates accelerate in growth and become statistically significant after WRAP participants reach the age of 65. When *APOE* is excluded, for the immediate learning composite score, the simple slope estimates of p-PRS under the endocytosis and APP metabolism pathways are only significant with cognitive decline once people reach age 80. For the delayed recall composite score, the simple slope estimate of overall PRS and p-PRSs under the cholesterol metabolism pathway become significant once people reach age 80 whereas the simple slope estimates of p-PRSs under the endocytosis and APP metabolism pathways become statistically significant once people reach age 75. For the executive function composite score, the simple slope estimates of overall PRS and p-PRS under the endocytosis pathway become significant once people reach age 75. For the PACC-3 score, the interaction patterns suggest that the adverse genetic effect starts to emerge once WRAP participants reach age 75 for the overall PRS and p-PRSs under the endocytosis and APP metabolism pathways.

**Figure 2.**
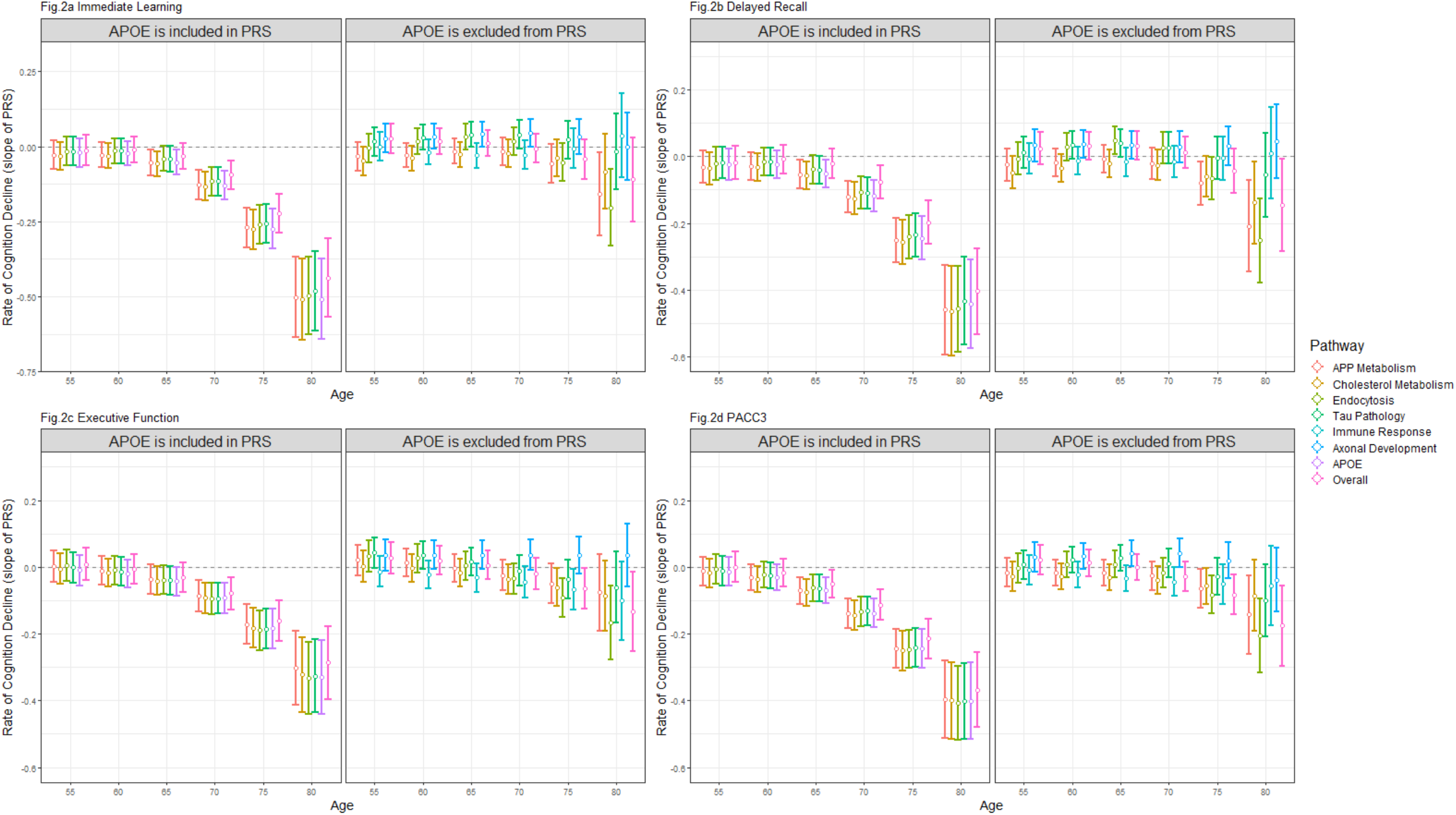
Model predicted simple slope of p-PRSs/PRS on cognition at different age with 95% confience interval in WRAP. presents the model predicted simple slope (based on the estimates) of p-PRSs/PRS on immediate learning (Figure 2a), delayed recall (Figure 2b), executive function (Figure 2c), and PACC3 (Figure 2d) at various age points in WRAP with 95% confidence intervals. Within each figure, the left panel depicts the simple slope estimates of p-PRSs/PRS, excluding *APOE* score, and the simple slope estimates of p-PRSs/PRS including *APOE* score are shown in the right panel. *APOE* is not theoretically affecting immune response and axonal development pathways, so the simple slope estimates of p-PRSs/PRS of these two pathways are only shown in the left panel. All association analyses are performed using the linear mixed effect model and adjusted for within-individual and within-family correlation. In addition to p-PRSs/PRS, age (linear, quadratic, and cubic), and their interactions, additional covariates include gender, education years, practice effect, and the first five genetic principal components of ancestry. Simple slope estimates were calculated based on parameters from the linear mixed effect model.

We used a reduced set of WRAP participants who have complete data in all PRS/p-PRSs to compare the performance of each PRS/p-PRS in explaining the amount of variation in the overall and domain-specific cognitive composite score, as measured by 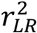 and presented in supplementary table 1. Consistent with Darst (2017), the largest 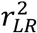 for a single PRS/p-PRS is about 0.2% when *APOE* is included and 0.1% when *APOE* is excluded, which indicates that almost none of the model variance was explained by any of the single PRS/p-PRS. When the interaction between age and PRS/p-PRS was included, an additional 1% of the model variation was explained for all pathways when *APOE* was included, but the additional gain in explained model variation substantially decreased after *APOE* was excluded.

### CSF biomarker outcomes

Like the cognitive outcomes, significant interactions between the PRSs of all pathways and linear age were observed for all Aβ outcomes when *APOE* was included in the PRS (Figure 1b; full regression outputs are available upon request). However, for P-tau, significant interactions were only observed between age and overall PRS, as well as p-PRSs under APP metabolism, cholesterol metabolism, and endocytosis pathways. For T-tau, significant interactions were only observed between age and overall PRS, as well as p-PRS under the endocytosis pathway. When *APOE* was excluded from the PRS, significant interactions were observed between age and overall PRS, as well as p-PRSs under APP metabolism, cholesterol metabolism, and immune response pathway for Aβ 42; between age and p-PRSs under APP metabolism and cholesterol metabolism pathways for Aβ 42/40; between age and overall PRS, as well as p-PRSs under APP metabolism, cholesterol metabolism, endocytosis, and immune response pathways for P-tau; and between age and overall PRS, as well as p-PRSs under APP metabolism, endocytosis, and the immune response pathway for T-tau.

Figure 3 presents the simple slope estimates for annual biomarker change by a one standard deviation change in genetic risk of PRS/p-PRS and by age in five-year increments between 50 and 80 years old for WRAP participants. When *APOE* is included, on average, significant simple slope estimates of p-PRSs under all pathways for Aβ42 emerge at around age 60, and the absolute effect sizes of these estimates increase with age. The adverse effect of p-PRSs on the Aβ42/40 ratio showed a pattern similar to Aβ42, but the model parameters predict that significant p-PRS/PRS-related differences in Aβ42/40 emerge at approximately age 55. For P-tau and T-tau, even though we observed a similar age-related genetic risk for all pathways when *APOE* was included in the p-PRS/PRS, the model parameters predict that significant PRS/PRS-related differences emerge a decade later than that for the Aβ outcome. When *APOE* was excluded, significant simple slope estimates of annual change for p-PRSs on beta-amyloid biomarkers still existed for some pathways but were less obvious, becoming statistically significant at an older age and with smaller absolute effect size. Specifically, model parameters predict that significant p-PRS/PRS-related differences in Aβ42 will emerge at age 70 for APP metabolism and cholesterol metabolism, and at age 75 for the immune response pathway. Similar findings were observed for the Aβ42/40 ratio, but the simple slope estimates of the p-PRSs under APP metabolism, cholesterol metabolism, and immune response become significant about 5∼10 years earlier than those predicted for the change of Aβ42, and model parameters predict that significant overall PRS-related differences in Aβ42/40 emerge at age 65. Surprisingly, the removal of *APOE* from p-PRSs does not affect the model-predicted, simple-slope estimates on tau for the pathways that should theoretically include *APOE*. For P-tau, after the removal of *APOE*, significant simple slope estimates for the p-PRSs appear once people reach age 65 for APP metabolism and the cholesterol metabolism pathway; at age 70 for the overall PRS and p-PRS under the endocytosis pathway; and at age 80 for the immune response p-PRS. Statistically significant simple slope estimates of the p-PRSs under APP metabolism and cholesterol metabolism on T-tau appear once people reach the age of around 65, whereas the model predicted simple slope estimates for p-PRSs under endocytosis and overall PRS are positively associated with a T-tau increase once people reach age 75.

**Figure 3.**
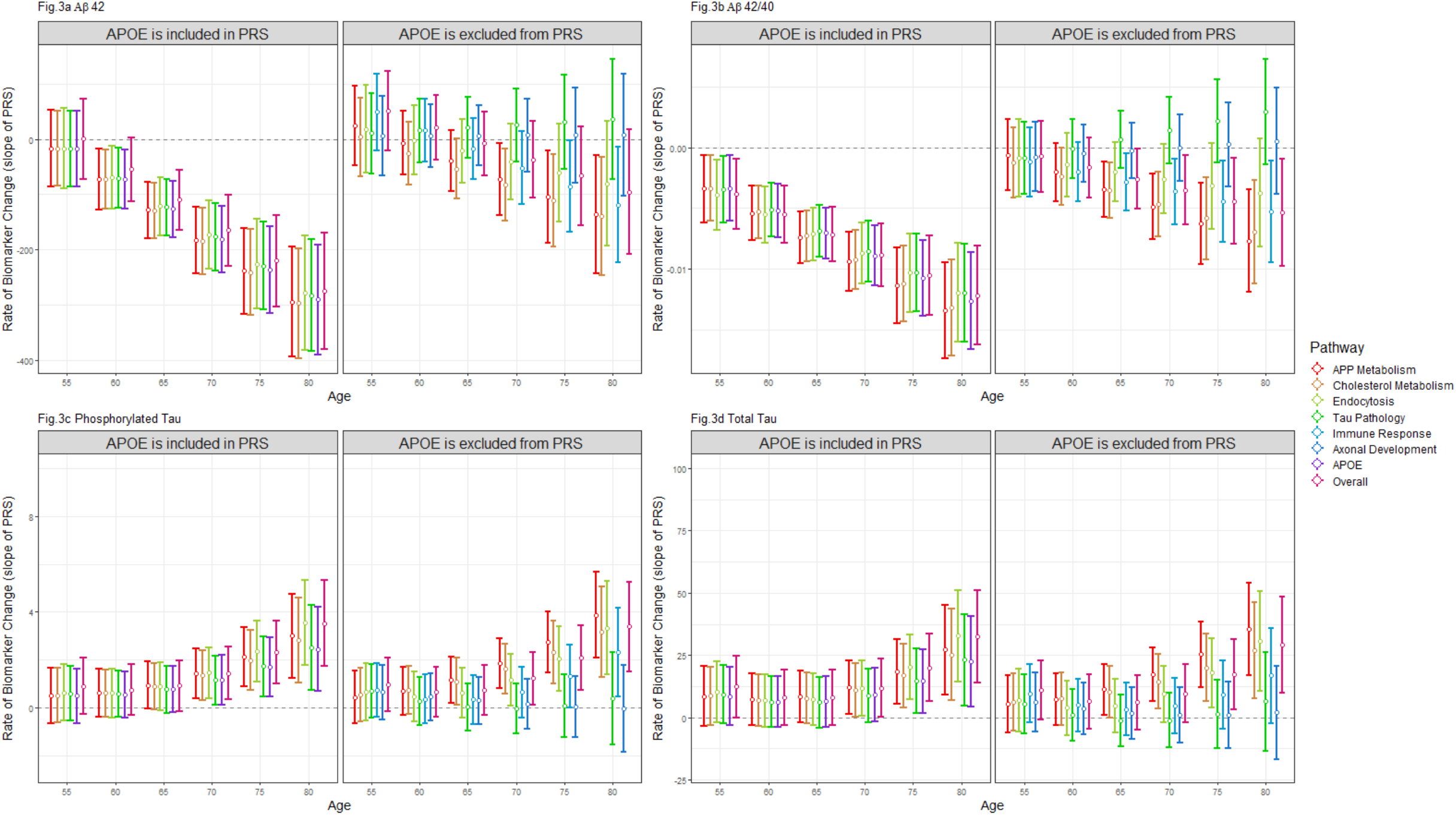
Model predicted simple slope of p-PRSs/PRS on biomarker at different age with 95% confience interval in WRAP. presents the model predicted simple slope (based on the estimates) of p-PRSs/PRS on beta-amyloid 42 (Figure 3a), beta-amyloid 42/40 ratio (Figure 3b), phosphorylated tau (Figure 3c), and total tau (Figure 3d) at various age points in WRAP with 95% confidence interval. Within each figure, the left panel depicts the simple slope estimates of p-PRSs/PRS, excluding *APOE* score, and the simple slope estinates of p-PRSs/PRS including *APOE* score are shown in the right panel. *APOE* is not theoretically affecting immune response and axonal development pathways, so the simple slope estimates of p-PRSs/PRS of these two pathways are only shown in the left panel. All association analyses are performed using the linear mixed effect model and adjusted for within-individual and within-family correlation. Spaghetti plots determine the functional form of age for all biomarker analyses. In addition to p-PRSs/PRS, age, and their interactions, additional covariates include gender, education years, and the first five genetic principal components of ancestry. Simple slope estimates were calculated based on parameters from the linear mixed effect model.

We used a reduced set of WRAP participants who have complete data in all PRS/p-PRSs to compare the performance of each PRS/p-PRS in explaining the amount of variation in the LOAD-related biomarkers, as measured by 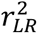 and presented in supplementary table 2. When *APOE* is included, single p-PRS/PRS can explain on average a 3∼4% variance in Aβ42, 7-8% variance in Aβ42/40, and 1% variance in T-tau and P-tau. Adding an interaction between PRS/p-PRS and age resulted in an additional 3∼4%, 1∼3%, 1-2% gain in the variance explained for Aβ42, Aβ42/40, T-tau and P-tau, respectively. For beta-amyloid outcomes, the gain in variance explained by addition of the p-PRS/PRS and the additional gain in model variation explained by the interaction between age and PRS/p-PRSs was substantially decreased after *APOE* was excluded. However, the removal of *APOE* from p-PRS/PRS does not substantially affect the additional variance explained by the PRS and interaction term for the tau outcomes.

### Replication analysis

We used the AHC combined sample from the Wisconsin ADRC to replicate our findings in WRAP. Table 2 details the Wisconsin ADRC participant characteristics for the cognition analysis. The mean baseline age for the AHC cohort is about 60. The mean education is just over 16 years. About 35% of participants in the AHC cohort are males, and 70% have a family history of AD. The basic characteristics are similar between the WRAP and AHC cohorts. Similar characteristics were observed in the biomarker samples.

**Table 2.** Participant characteristics for the Wisconsin ADRC

| Variable | Wisconsin ADRC AHC |  |
| --- | --- | --- |
|  | PACC3-TMT (N=427) | Biomarker Biomarker (N=259) |
| Baseline age | 59.95 (8.26) | 60.53 (8.08) |
| Education (years) | 16.35 (2.46) | 16.20 (2.39) |
| Gender (male) | 149 (35%) | 80 (31%) |
| Family history of AD | 299 (70%) | 192 (74%) |
| **Max visits** |  |  |
| 1 | 31 (7%) | 203 (78%) |
| 2 | 56 (13%) | 34 (13%) |
| 3 | 60 (14%) | 6 (2%) |
| 4 | 34 (8%) | 11 (4%) |
| 5 | 246 (58%) | 3 (1%) |
| 6 |  | 1 (0.4%) |
| 7 |  | 1 (0.4%) |
| **APOE genotype** |  |  |
| e2/e2 | 1 (0.2%) | 1 (0.4%) |
| e2/e3 | 49 (11%) | 30 (12%) |
| e3/e3 | 219 (51%) | 132 (51%) |
| e2/e4 | 14 (3%) | 7 (3%) |
| e3/e4 | 124 (29%) | 76 (29%) |
| e4/e4 | 20 (5%) | 13 (5%) |
| CSF Aβ42 (pg/mL) | N/A | 957.40 (381.79) |
| CSF Aβ42/40 | N/A | 0.07 (0.01) |
| CSF T-tau (pg/mL) | N/A | 194.47 (73.58) |
| CSF P-tau (pg/mL) | N/A | 17.00 (6.91) |

Mean (SD); n (%); AHC: all healthy controls; Aβ : Beta-amyloid; P-tau: phosphorylated Tau; T-tau: total Tau.

For the cognition analysis, the correlation between PACC-3 and PACC-3-TMT is about 0.93 in the WRAP sample. The results from the Wisconsin ADRC are mostly consistent with the WRAP findings in terms of the genetic risk on cognition being stronger in older ages, but were less obvious (Figure 1c). Specifically, when *APOE* was included in the p-PRS/PRS, statistically significant interactions were observed between polynomial age and p-PRS under the tau pathology pathway only. When *APOE* was excluded, significant interactions were observed between polynomial age and p-PRS under the APP metabolism and cholesterol metabolism pathways.

When *APOE* was included, the model-predicted simple slope estimates (supplementary figure 2) for the overall PRS and p-PRS under tau pathology and endocytosis pathways become significant at age 85. The simple slope estimates of *APOE* are not statistically different from zero in any age range. When *APOE* is excluded, the model parameters predict that significant p-PRS-related differences in PACC-3-TMT emerge after age 70 for both APP metabolism and cholesterol metabolism pathways. A reduced set of Wisconsin ADRC participants who have complete data in all PRS/p-PRSs were used to compare the performance of each PRS/p-PRS (supplementary table 3). Similar to the WRAP findings, the largest 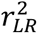 for a single PRS/p-PRS is about 0.2% in the AHC sample. The largest 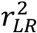 for PRS/p-PRSs with and without *APOE* increased by 0.4% and 0.7% with the addition of the interaction between PRS/p-PRSs and age, respectively.

Significant interactions between age and genetic risk were also observed in the biomarker analysis (Figure 1d). Specifically, when *APOE* is included in the PRS, significant interactions are observed between age and the p-PRSs of APP metabolism and cholesterol metabolism for Aβ42; between age and the p-PRSs/PRS of all pathways for Aβ42/40 and P-tau; and between age and overall PRS, as well as p-PRSs under endocytosis and tau pathology pathways for T-tau. When *APOE* was excluded, significant interactions were observed between age and p-PRS under the cholesterol metabolism pathway for Aβ42/40; between age and overall PRS, as well as p-PRSs under endocytosis and tau pathology pathways for both P-tau and T-tau. No significant interactions were observed for Aβ42, even though suggestive evidence of interactions were observed between age and p-PRSs under APP metabolism (*p*=0.07) and cholesterol metabolism pathways (*p*=0.06).

When *APOE* was included, on average, the model predicted the simple slope estimates of overall PRS and p-PRS under all pathways, becoming significant once people reach age 55, and the absolute effect size increases with age. Results for the Aβ42/40 ratio are very consistent with the WRAP findings. When *APOE* is included, the model parameters predict that significant p-PRS/PRS-related differences in Aβ42/40 emerge at approximately age 55, and the interaction patterns are similar to that of *APOE* alone. Results for T-tau and P-tau are also very similar to the WRAP findings when *APOE* is included. Significant simple slope estimates of the PRS/p-PRSs of all pathways start to appear once people reach age 65 for both P-tau and T-tau. When *APOE* is excluded and for Aβ42, similar to WRAP findings, only the simple slope estimates for p-PRSs under the APP metabolism and cholesterol metabolism pathways become significant once people reach age 65. For the Aβ42/40 ratio, when *APOE* is excluded, the simple slope estimate of p-PRS under the cholesterol metabolism pathway becomes significant at age 65, but the absolute effect size is smaller than that of the *APOE* score. For P-tau, significant simple slope estimates of the p-PRS start to appear at age 70 for the axonal development pathway and at age 75 for endocytosis, tau pathology, and overall PRS. The model parameters predict that significant differences in T-tau emerge at approximately 75 for tau pathology and axon development pathways and at age 80 for endocytosis and overall PRS. A reduced set of Wisconsin ADRC participants with complete data in all PRS/p-PRSs was used to compare the performance of each PRS/p-PRS (supplementary table 3). When *APOE* is included, the variance explained by a single PRS is about 3% for Aβ42, 7% for Aβ42/40, 1% for P-tau, and less than 1% for T-tau. The additional interaction between age and p-PRSs contributes to the added variance explained in Aβ42, Aβ42/40, T-tau, and P-tau by about 1%, 2%, 2%, and 1.5%, respectively. When *APOE* was excluded, p-PRSs and age–PRS interaction under all pathways contributed substantially less variance, as shown using Aβ42 and Aβ42/40 ratios, which is consistent with the WRAP findings. For tau-related outcomes, when *APOE* is excluded, the performance of p-PRSs/PRS and the interaction between p-PRSs/PRS and age under most pathways (except for endocytosis) deteriorated.

## Discussion

In the present study, we updated findings from Darst et al. and investigated the potential of pathway-specific PRSs in predicting rate of change in cognitive function and biomarkers of beta-amyloid deposition, tau pathology, and neurodegeneration among asymptomatic individuals in WRAP [19]. With five additional years of data collection, GWAS summary statistics with a larger sample size, our comprehensive variant-pathway mapping method, and the inclusion of an age-interaction effect, we found p-PRSs and the overall PRS can predict preclinical changes in cognition and biomarkers. The effects of p-PRSs/PRS on rate of change in cognition, beta-amyloid, and tau outcomes are related to age and appear earlier in the lifespan when *APOE* is included in these risk scores compared to when *APOE* is excluded. Consistent with Darst et al., *APOE* appears to drive much of the strength of the p-PRSs for APP metabolism, cholesterol metabolism, endocytosis, tau pathology, and the overall PRS on rate of change in cognition and beta-amyloid outcomes. However, we did not observe a similar *APOE*-driven effect trend when applying p-PRSs/PRS to predicting tau outcomes.

Results are mixed for current p-PRS studies on LOAD risk, cognitive decline, and biomarker variation. This is partially due to the discrepancy in the sample characteristics across different studies and outcomes being investigated, but also due to the methodology of attributing specific genetic variants to its corresponding biological pathway. Most existing studies on p-PRSs have been based entirely on limited or even single-review papers and bioinformatic databases to map a specific genetic variant to a neurobiological pathway. However, the genetic functions of a specific variant have not been consistently defined across the literature, and the functional annotation of genes might differ across various databases [22]. This creates uncertainties in the accuracy of constructing p-PRSs and creates the possibility that the same pathway various studies explored might not be comparable and a specific pathway might not comprehensively reflect the underlying biological mechanism that it intends to represent. A recent study on p-PRSs proposed a novel approach to construct p-PRSs by including a multiplicative factor that represents the degree of involvement of a given genetic variant in the preselected pathways in calculating p-PRSs to allow for uncertainty in gene and pathway assignment [22]. Even though this approach overcomes some limitations that the traditional p-PRSs studies may encounter when constructing p-PRSs, the accuracy of the p-PRSs under this approach (overweight or underweight of a particular variant) and to what extent the resulting p-PRSs reflect the underlying biological mechanism are still unknown. In our study, we combined the merits of these two approaches and proposed a conservative but comprehensive variant-pathway mapping method via only matching variants and corresponding pathways for which we have good confidence of an “actually existing” biological relationship, after widely referring to the literature published within recent years.

Although genetics play a large role in the development and expression of LOAD, the complex relationships in the etiology between age, *APOE*, and non-*APOE* p-PRSs/PRS are not generally considered[43]. Our study shows the risk of p-PRSs/PRS on rate of change in cognition, beta-amyloid, and tau are age related in both WRAP and the Wisconsin ADRC, regardless of including *APOE*. However, the adverse effects of p-PRSs/PRS appear earlier in the lifespan when *APOE* is included in these risk scores compared to when *APOE* is excluded, with the exception of tau outcomes. In addition, including the interaction between p-PRSs/PRS and age in the model led to the model explaining additional variance. Our findings are consistent with a recent study that leveraged Alzheimer’s Disease Neuroimaging Initiative (ADNI and UK Biobank (UKBB) samples and concluded both *APOE* and PRS predicted AD risk and presented age-related effects, but the effects of *APOE* were stronger in younger groups (age <80)[44]. Zimmerman et al. examined the age-related genetic effect of *APOE* and PRS in UKBB. Similar to our findings, they reported that an AD PRS modified the association between age and cognition, that *APOE* ε4 allele carriers experienced earlier cognitive decline than non-carriers did, and that models using the PRS that excluded *APOE* ε4 had attenuated and later modification of age associations compared to when *APOE* was included in the PRS [45]. Our study also demonstrated a pattern in the timing of the earliest detectable genetic effect on rate of change in beta-amyloid (age ∼55), tau (age ∼65), and cognition (age ∼65–70) that aligns with findings from Hanseeuw et al. [46]. This pattern also explained that the reason for observing significant associations between p-PRSs/PRS and beta-amyloid outcomes in Darst et al., but not for tau and cognition outcomes is that beta-amyloid accumulation occurs at a younger age than variation in tau and cognition, and the sample analyzed in Darst et al. was too young to detect polygenic effects on tau and cognition.

Pathway-specific PRS could also predict earlier changes in AD-related outcomes than the overall PRS could, especially for beta-amyloid outcomes and when *APOE* is excluded from the risk score. Our results demonstrate when *APOE* is excluded, p-PRSs for APP and cholesterol pathway can predict changes in Aβ42, which is about 15 years earlier than the overall PRS, and this finding was replicated in the Wisconsin ADRC. Even though p-PRSs for APP and cholesterol metabolism pathways also show potential for predicting earlier changes in Aβ42/40 ratio compared to the overall PRS, only the finding for the cholesterol metabolism pathway was replicated in the Wisconsin ADRC. Our results also show p-PRSs under certain pathways can predict adverse change in tau and cognition outcomes earlier than the overall PRS in WRAP can, but these findings were not fully replicated in the Wisconsin ADRC, which may warrant further investigation as longitudinal data focusing on the preclinical stage of AD with a larger sample size become available.

One finding – that the tau pathology PRS is not predictive of tau outcomes after the exclusion of *APOE* in WRAP, but is predictive of both P-tau and T-tau in the Wisconsin ADRC - may require further investigation once we have a larger sample size and longer follow-up time. There are two possible explanations for the discrepancies in the effect of the tau pathology PRS between WRAP and the Wisconsin ADRC. First, the genes known to be related to the tau pathology pathway may not be well established and may not fully reflect the biological pathway of tau since only three SNPs in addition to *APOE* were included in the tau pathology pathway and all these SNPs overlapped with the SNPs included in the other disease-related pathways (Supplemental Figure 1). Second, the AHC sample that was extracted from the Wisconsin ADRC includes a sample of older healthy controls (enrollment age >= 65) and tau-levels are higher in the older age groups [47].

The present study has limitations. First, although we tend to match genetic variants and biological pathways for which we have good confidence of a biological variant-pathway relationship, our variant-pathway mapping method is conservative and may not comprehensively reflect the genetic role in a specific disease pathway. Our results’ accuracy is subject to the knowledge of biological function of genes and pathways at the time of performing this study. It would be crucial to modify the variant-pathway mapping as additional knowledge becomes available. Second, we only considered the significant variants as identified from the most recent IGAP case-control GWAS meta-analyses as the weight to construct pathway-specific PRSs; however, a larger panel of SNPs from the recent GWAS by proxy (GWAX) and a combined study of GWAS and GWAX may provide additional insights into the variant-pathway mapping [48,49]. This was not considered in the current study. Third, results from the AHC sample extracted from the Wisconsin ADRC are not absolutely comparable with the WRAP findings because the AHC sample was constructed based on two separate cohorts with different characteristics (e.g., age) Additional replication analyses may be carried out once the data for an older IMPACT cohort (more like WRAP) become available.

In conclusion, in addition to *APOE*, the pathway-specific PRSs can predict age dependent changes in beta-amyloid, tau, and cognition. Once validated, they could be used to identify individuals with an elevated genetic risk of accumulating beta-amyloid and tau, long before the onset of clinical symptoms. This information could be useful for selection of high risk participants for clinical trials and, as effective therapeutic targets further develop, p-PRSs could be used to determine an individual’s risk for accumulating amyloid and the predicted age of onset so that resources could be used effectively in screening individuals for amyloid accumulation with more expensive and invasive, but accurate tests. This idea is being explored in other diseases, such as breast cancer[50].

## Data Availability

The data supporting the findings of this study are available on request from the corresponding author. The data are not publicly available due to privacy or ethical restrictions.

## Acknowledgements

The authors especially thank the WRAP and Wisconsin ADRC participants and staff for their contributions to the studies. Without their efforts, this research would not be possible. This study was supported by the National Institutes of Health (NIH) grants R01AG27161 (Wisconsin Registry for Alzheimer Prevention: Biomarkers of Preclinical AD), R01AG054047 (Genomic and Metabolomic Data Integration in a Longitudinal Cohort at Risk for Alzheimer’s Disease), R21AG067092 (Identifying Metabolomic Risk Factors in Plasma and Cerebrospinal Fluid for Alzheimer’s Disease), and P30AG062715 (Wisconsin Alzheimer’s Disease Research Center Grant)], the Helen Bader Foundation, Northwestern Mutual Foundation, Extendicare Foundation, State of Wisconsin, the Clinical and Translational Science Award (CTSA) program through the NIH National Center for Advancing Translational Sciences (NCATS) grant [UL1TR000427]. Computational resources were supported by core grants to the Center for Demography and Ecology [P2CHD047873] and the Center for Demography of Health and Aging [P30AG017266]. Author YD was supported by the Biology of Aging and Age-Related Diseases training grant T32 AG000213-28 from the National Institute on Aging. HZ is a Wallenberg Scholar supported by grants from the Swedish Research Council (#2018-02532), the European Research Council (#681712 and #101053962), Swedish State Support for Clinical Research (#ALFGBG-71320), the Alzheimer Drug Discovery Foundation (ADDF), USA (#201809-2016862), the AD Strategic Fund and the Alzheimer’s Association (#ADSF-21-831376-C, #ADSF-21-831381-C, and #ADSF-21-831377-C), the Bluefield Project, the Olav Thon Foundation, the Erling-Persson Family Foundation, Stiftelsen för Gamla Tjänarinnor, Hjärnfonden, Sweden (#FO2022-0270), the European Union’s Horizon 2020 research and innovation programme under the Marie Skłodowska-Curie grant agreement No 860197 (MIRIADE), the European Union Joint Programme – Neurodegenerative Disease Research (JPND2021-00694), and the UK Dementia Research Institute at UCL (UKDRI-1003). KB is supported by the Swedish Research Council (#2017-00915), ADDF, USA [#RDAPB-201809-2016615], the Swedish Alzheimer Foundation [#AF-742881], Hjärnfonden, Sweden [#FO2017-0243], the Swedish state under the agreement between the Swedish government and the County Councils, the ALF-agreement [#ALFGBG-715986], and European Union Joint Program for Neurodegenerative Disorders [JPND2019-466-236].

Thank you to Roche Diagnostics International Ltd for providing the NeuroToolKit robust prototype assays s for this study. The Roche NeuroToolKit is a panel of exploratory prototype assays designed to robustly evaluate biomarkers associated with key pathologic events characteristic of AD and other neurological disorders, used for research purposes only and not approved for clinical use in the U.S..

