## Supplementary tables and figures for "Effect of Pathway-specific Polygenic Risk Scores for Alzheimer’s Disease (AD) on Rate of Change in Cognitive Function and AD-related Biomarkers among Asymptomatic Individuals"

Running Title: AD Pathway-specific Polygenic Risk Score

Yuexuan **Xu**[a] Eva **Vasiljevic**[a,n], Yuetiva K. **Deming**[a,b,c], Erin M. **Jonaitis**[c], Rebecca L. **Koscik**[b,c,d], Carol A. **Van Hulle**[b,d], Qiongshi **Lu**[e], Margherita **Carboni**[f], Gwendlyn **Kollmorgen**[g], Norbert **Wild**[g], Cynthia M. **Carlsson**[b,d,h], Sterling C. **Johnson**[b,d], Henrik **Zetterberg**[i,j,k,l,m], Kaj **Blennow**[i,j], Corinne D. **Engelman**[a]

[a]Department of Population Health Sciences, School of Medicine and Public Health, University of Wisconsin-Madison, WI, USA

[b]Department of Medicine, School of Medicine and Public Health, University of Wisconsin-Madison, WI, USA

[c]Wisconsin Alzheimer's Institute, University of Wisconsin-Madison, WI, USA

[d]Wisconsin Alzheimer's Disease Research Center, University of Wisconsin-Madison, WI, USA

[e]Department of Biostatistics and Medical Informatics, School of Medicine and Public Health, University of Wisconsin-Madison, WI, USA

[f]Roche Diagnostics International Ltd, Rotkreuz, Switzerland

[g]Roche Diagnostics GmbH, Penzberg, Germany

[h]Geriatric Research Education and Clinical Center, Wm. S. Middleton Memorial VA Hospital, Madison, WI, USA

[i]Department of Psychiatry and Neurochemistry, Institute of Neuroscience and Physiology, The Sahlgrenska Academy at University of Gothenburg, Mölndal, Sweden

[j]Clinical Neurochemistry Laboratory, Sahlgrenska University Hospital, Mölndal, Sweden

[k]UK Dementia Research Institute at UCL, London, UK

[l]Department of Neurodegenerative Disease, UCL Institute of Neurology, London, UK

[m]Hong Kong Center for Neurodegenerative Diseases, Hong Kong, China

[n]Center for Demography of Health and Aging, University of Wisconsin-Madison, WI, USA

**Correspondence:**

Corinne D. Engelman, MSPH, PhD

Department of Population Health Sciences,

University of Wisconsin - Madison

610 Walnut Street, 1007A WARF

Madison, WI 53726-2397

### Supplementary files

#### Supplementary methods

*Model setting*

For each PRSs we have investigated in our study, we fitted the following linear mixed effect model.

$$y_{it} = \beta_1 PRS + \beta_2 Age_{it} + \beta_3 Age_{it}^2 + \beta_4 Age_{it}^3 + \beta_5 PRS * Age_{it} + \beta_6 PRS * Age_{it}^2 + \beta_7 PRS * Age_{it}^3 + \sum_{j=1}^{m} \beta_{wj} W_{ij} + \sum_{k=1}^{p} \beta_{xk} X_{itk} + Z_u + \epsilon$$

Where y is the outcome being studied, PRS is the overall/pathway specific PRS, age is the continuous age at each visit, W is a vector of time-invariant predictors (gender, education, principal components), and X is a set of time-varying predictors (practice effect). Z is a vector of random effects and e is the residual vector.

*Likelihood ratio tests*

To compute the overall significance level of the interaction term (linear, quadratic, and cubic interaction term), we used the likelihood ratio test (LRT) and test the null hypothesis that the coefficients for linear, quadratic, and cubic interaction terms are simultaneously zero (degree of freedom = difference between number of parameters in the full model vs nested model).

*Simple slope analysis*

Upon the discovery of significant interactions, we probe the nature of the interaction effects by using the simple slope (conditional effect) approach. The conditional effects of PRS (simple slope) based on different value of age is calculated using the following equation (for an example please see supplementary excel table).

$$Slope(PRS) = PRS(\beta_1 + \beta_5 Age_{it} + \beta_6 Age_{it}^2 + \beta_7 Age_{it}^3)$$

Supplementary table 1. Incremental likelihood ratio R-squared ($r_{LR}^2$ ) for each cognitive outcome model based on a reduced subset of WRAP participants who were not missing PRS measures (n=1073).

| | **APP** | | **Immune** | | **Cholesterol** | | **Endocytosis** | | **Tau** | | **Axon** | | ***APOE*** | | **Overall** | |
|---|---|---|---|---|---|---|---|---|---|---|---|---|---|---|---|---|
| ***Outcome*** | ***PRS*** | ***Interaction*** | ***PRS*** | ***Interaction*** | ***PRS*** | ***Interaction*** | ***PRS*** | ***Interaction*** | ***PRS*** | ***Interaction*** | ***PRS*** | ***Interaction*** | ***APOE Score*** | ***Interaction*** | ***PRS*** | ***Interaction*** |
| | | | | | | | | ***APOE* included** | | | | | | | | |
| Delayed Recall | **0.002** | 0.009 | / | / | **0.002** | 0.009 | 0.001 | **0.010** | 0.001 | 0.008 | / | / | 0.001 | 0.008 | 0.001 | 0.009 |
| Executive Function | **0.001** | 0.007 | / | / | **0.001** | 0.007 | 0.000 | **0.008** | 0.000 | 0.007 | / | / | **0.001** | 0.006 | 0.000 | 0.007 |
| Immediate Learning | **0.002** | **0.011** | / | / | **0.002** | 0.010 | 0.001 | **0.011** | 0.001 | 0.010 | / | / | **0.002** | 0.010 | 0.001 | 0.010 |
| PACC3 | **0.002** | 0.010 | / | / | **0.002** | 0.010 | **0.002** | **0.011** | 0.001 | 0.010 | / | / | **0.002** | 0.010 | 0.001 | **0.011** |
| | | | | | | | | ***APOE* excluded** | | | | | | | | |
| Delayed Recall | 0.000 | 0.002 | 0.000 | 0.000 | **0.001** | 0.001 | 0.000 | **0.005** | 0.000 | 0.001 | **0.001** | 0.000 | **/** | / | 0.000 | 0.001 |
| Executive Function | 0.000 | 0.001 | 0.000 | 0.001 | 0.000 | 0.001 | 0.000 | **0.005** | 0.000 | 0.002 | 0.000 | 0.000 | / | / | 0.000 | 0.002 |
| Immediate Learning | 0.000 | 0.001 | 0.000 | 0.001 | **0.001** | 0.000 | 0.000 | **0.002** | 0.000 | 0.000 | **0.001** | 0.000 | / | / | 0.000 | 0.001 |
| PACC3 | 0.000 | 0.001 | 0.000 | 0.001 | **0.001** | 0.000 | 0.000 | **0.004** | 0.000 | 0.001 | **0.001** | 0.001 | / | / | 0.000 | 0.003 |

The incremental R-squared for each PRS/p-PRS is calculated as the difference in model likelihood ratio R-squared between the PRS model and covariate model (i.e., the covariate model with PRS/p-PRS and without PRS/p-PRS). The incremental R-squared for the interaction terms is calculated as the difference in model likelihood ratio R-squared between the interaction model and the PRS model (i.e., the PRS model with and without interaction terms). The covariate model below is the base model for comparison; there are no results for this model and it is not included in the table.

Covariate model: age + sex + education + practice effects + principal components

PRS model: age + sex + education + practice effects + principal components + **PRS**

Interaction model: age + sex + education + practice effects + principal components + **PRS + PRS × age**

*Age form (as determined by previous literature): cognition (linear, quadratic, and cubic terms)

Bolded values are the highest incremental R-squared for each outcome for the PRS model and the interaction model. For example, for Delayed Recall when *APOE* is included, the APP and Cholesterol PRS have the highest incremental R-squared (they explain the highest percent increase in variance explained compared to the covariate model) and the Endocytosis PRS*age interaction has the highest incremental R-squared compared to the PRS model.

APP metabolism pathway: *CLU, SORL1, ABCA7, PICALM, ADAM10, APOE*
Cholesterol metabolism pathway: *CLU, SORL1, ABCA7, APOE*
Endocytosis pathway: *SORL1, ABCA7, PICALM, BIN1, CD2AP, PTK2B, FERMT2, SLC24A4, APOE*
Tau pathway: *BIN1, FERMT2, CASS4, APOE*
Immune response: *CLU, ABCA7, CR1, INPP5D, HLA-DRB1, TREM2, EPHA1, MS4A6A, CD33, MEF2C*
Axonal development: *EPHA1, FERMT2, CASS4, SPI1, NME8*

Supplementary table 2. Incremental likelihood ratio R-squared ($r_{LR}^2$ ) for each biomarker outcome model based on a reduced subset of WRAP participants who were not missing PRS measures (n=172)

| **Outcome** | **APP** | | **Immune** | | **Cholesterol** | | **Endocytosis** | | **Tau** | | **Axon** | | ***APOE*** | | **Overall** | |
|---|---|---|---|---|---|---|---|---|---|---|---|---|---|---|---|---|
| | **PRS** | **Interaction** | **PRS** | **Interaction** | **PRS** | **Interaction** | **PRS** | **Interaction** | **PRS** | **Interaction** | **PRS** | **Interaction** | **APOE Score** | **Interaction** | **PRS** | **Interaction** |
| ***APOE* included** | | | | | | | | | | | | | | | | |
| Aβ42 | 0.039 | 0.035 | / | / | **0.041** | 0.035 | 0.036 | 0.032 | 0.035 | 0.029 | / | / | 0.039 | 0.031 | 0.029 | **0.037** |
| Aβ42/40 | **0.079** | **0.024** | / | / | 0.078 | 0.022 | 0.073 | 0.018 | 0.066 | 0.016 | / | / | 0.071 | 0.019 | 0.072 | 0.022 |
| P-tau | 0.009 | 0.020 | / | / | 0.008 | 0.016 | 0.008 | 0.019 | 0.006 | 0.011 | / | / | 0.005 | 0.012 | **0.010** | **0.021** |
| T-tau | **0.009** | 0.017 | / | / | 0.007 | 0.014 | 0.007 | 0.018 | 0.005 | 0.013 | / | / | 0.005 | 0.012 | **0.009** | **0.021** |
| ***APOE* excluded** | | | | | | | | | | | | | | | | |
| Aβ42 | 0.001 | 0.007 | 0.000 | **0.010** | **0.004** | 0.006 | 0.000 | 0.003 | 0.000 | 0.000 | 0.000 | 0.000 | / | / | 0.000 | **0.010** |
| Aβ42/40 | 0.013 | **0.011** | 0.01 | 0.007 | **0.015** | 0.006 | 0.005 | 0.001 | 0.000 | 0.002 | 0.001 | 0.001 | / | / | 0.008 | 0.007 |
| P-tau | **0.013** | **0.036** | 0.003 | 0.017 | 0.010 | 0.014 | 0.006 | 0.019 | 0.001 | 0.004 | 0.002 | 0.001 | / | / | 0.008 | 0.022 |
| T-tau | **0.012** | **0.027** | 0.003 | 0.015 | 0.008 | 0.009 | 0.004 | 0.017 | 0.000 | 0.005 | 0.002 | 0.001 | / | / | 0.006 | 0.021 |

The incremental R-squared for each PRS/p-PRS is calculated as the difference in model likelihood ratio R-squared between the PRS model and covariate model (i.e., the covariate model with PRS/p-PRS and without PRS/p-PRS). The incremental R-squared for the interaction terms is calculated as the difference in model likelihood ratio R-squared between the interaction model and the PRS model (i.e., the PRS model with and without interaction terms). The covariate model below is the base model for comparison; there are no results for this model and it is not included in the table.

Covariate model: age + sex + education + practice effects + principal components

PRS model: age + sex + education + practice effects + principal components + **PRS**

Interaction model: age + sex + education + practice effects + principal components + **PRS + PRS × age**

*Age form (as determined by spaghetti plot): beta-amyloid outcomes (linear term); tau-outcomes (linear and quadratic terms)

Bolded values are the highest incremental R-squared for each outcome for the PRS model and the interaction model. For example, for Delayed Recall when *APOE* is included, the APP and Cholesterol PRS have the highest incremental R-squared (they explain the highest percent increase in variance explained compared to the covariate model) and the Endocytosis PRS*age interaction has the highest incremental R-squared compared to the PRS model.

APP metabolism pathway: *CLU, SORL1, ABCA7, PICALM, ADAM10, APOE*
Cholesterol metabolism pathway: *CLU, SORL1, ABCA7, APOE*
Endocytosis pathway: *SORL1, ABCA7, PICALM, BIN1, CD2AP, PTK2B, FERMT2, SLC24A4, APOE*
Tau pathway: *BIN1, FERMT2, CASS4, APOE*
Immune response: *CLU, ABCA7, CR1, INPP5D, HLA-DRB1, TREM2, EPHA1, MS4A6A, CD33, MEF2C*
Axonal development: *EPHA1, FERMT2, CASS4, SPI1, NME8*

Supplementary table 3. Incremental likelihood ratio R-squared ($r_{LR}^2$ ) for each cognitive and biomarker outcome model based on a reduced subset of Wisconsin ADRC participants who were not missing PRS measures (cognition sample: n=361, biomarker sample: n=216)

| Outcome | APP | | Immune | | Cholesterol | | Endocytosis | | Tau | | Axon | | APOE | | Overall | |
|---|---|---|---|---|---|---|---|---|---|---|---|---|---|---|---|---|
| | **PRS** | **Interaction** | **PRS** | **Interaction** | **PRS** | **Interaction** | **PRS** | **Interaction** | **PRS** | **Interaction** | **PRS** | **Interaction** | **Score** | **Interaction** | **PRS** | **Interaction** |
| **APOE included** | | | | | | | | | | | | | | | | |
| PACC3_TMT | **0.001** | 0.003 | **/** | / | **0.001** | 0.003 | **0.001** | **0.004** | **0.001** | **0.004** | **/** | / | **0.001** | 0.003 | 0.000 | 0.003 |
| Aβ42 | 0.031 | **0.010** | / | / | **0.033** | **0.010** | 0.031 | 0.004 | 0.024 | 0.003 | / | / | 0.027 | 0.004 | 0.028 | **0.010** |
| Aβ42/40 | 0.075 | 0.023 | / | / | **0.079** | 0.023 | 0.066 | 0.022 | 0.066 | 0.021 | / | / | 0.071 | 0.019 | 0.065 | **0.026** |
| P-tau | 0.009 | 0.020 | / | / | **0.010** | 0.022 | 0.007 | 0.023 | **0.010** | 0.022 | / | / | **0.010** | 0.020 | **0.010** | **0.027** |
| T-tau | 0.004 | 0.015 | / | / | 0.004 | 0.017 | 0.003 | 0.016 | **0.005** | 0.016 | / | / | **0.005** | 0.016 | 0.004 | **0.019** |
| **APOE excluded** | | | | | | | | | | | | | | | | |
| PACC3_TMT | 0.000 | 0.005 | **0.002** | 0.001 | 0.000 | **0.007** | 0.000 | 0.003 | 0.000 | 0.003 | **0.002** | 0.001 | / | / | **0.002** | 0.001 |
| Aβ42 | 0.005 | 0.007 | 0.000 | 0.002 | **0.009** | **0.010** | 0.003 | 0.000 | 0.002 | 0.001 | 0.000 | 0.001 | **/** | / | 0.000 | 0.000 |
| Aβ42/40 | 0.001 | 0.001 | 0.000 | 0.005 | **0.006** | **0.007** | 0.000 | 0.000 | 0.002 | 0.002 | 0.001 | 0.001 | / | / | 0.000 | 0.002 |
| P-tau | 0.001 | 0.010 | **0.002** | 0.007 | 0.000 | 0.000 | **0.002** | 0.029 | 0.000 | **0.030** | **0.002** | 0.017 | / | / | 0.000 | 0.016 |
| T-tau | 0.004 | 0.010 | 0.000 | 0.006 | 0.001 | 0.000 | **0.005** | **0.035** | 0.000 | 0.029 | 0.001 | 0.013 | / | / | 0.001 | 0.017 |

The incremental R-squared for each PRS/p-PRS is calculated as the difference in model likelihood ratio R-squared between the PRS model and covariate model (i.e., the covariate model with PRS/p-PRS and without PRS/p-PRS). The incremental R-squared for the interaction terms is calculated as the difference in model likelihood ratio R-squared between the interaction model and the PRS model (i.e., the PRS model with and without interaction terms). The covariate model below is the base model for comparison; there are no results for this model and it is not included in the table.

Covariate model: age + sex + education + practice effects + principal components

PRS model: age + sex + education + practice effects + principal components + **PRS**

Interaction model: age + sex + education + practice effects + principal components + **PRS + PRS × age**

*Age form (as determined by previous analysis and spaghetti plot): cognition (linear, quadratic, and cubic terms); beta-amyloid outcomes (linear term); tau-outcomes (linear and quadratic terms)

Bolded values are the highest incremental R-squared for each outcome for the PRS model and the interaction model. For example, for Delayed Recall when *APOE* is included, the APP and Cholesterol PRS have the highest incremental R-squared (they explain the highest percent increase in variance explained compared to the covariate model) and the Endocytosis PRS*age interaction has the highest incremental R-squared compared to the PRS model.

APP metabolism pathway: *CLU, SORL1, ABCA7, PICALM, APOE*
Cholesterol metabolism pathway: *CLU, SORL1, ABCA7, APOE*
Endocytosis pathway: *SORL1, ABCA7, PICALM, BIN1, CD2AP, PTK2B, FERMT2, SLC24A4, APOE*
Tau pathway: *BIN1, FERMT2, CASS4, APOE*
Immune response: *CLU, ABCA7, CR1, INPP5D, HLA-DRB1, TREM2, EPHA1, MS4A6A, CD33, MEF2C*
Axonal development: *EPHA1, FERMT2, CASS4, SPI1, NME8*

#### Supplementary Figures

**Supplementary figure 1. Venn diagram of the gene-pathway mapping**

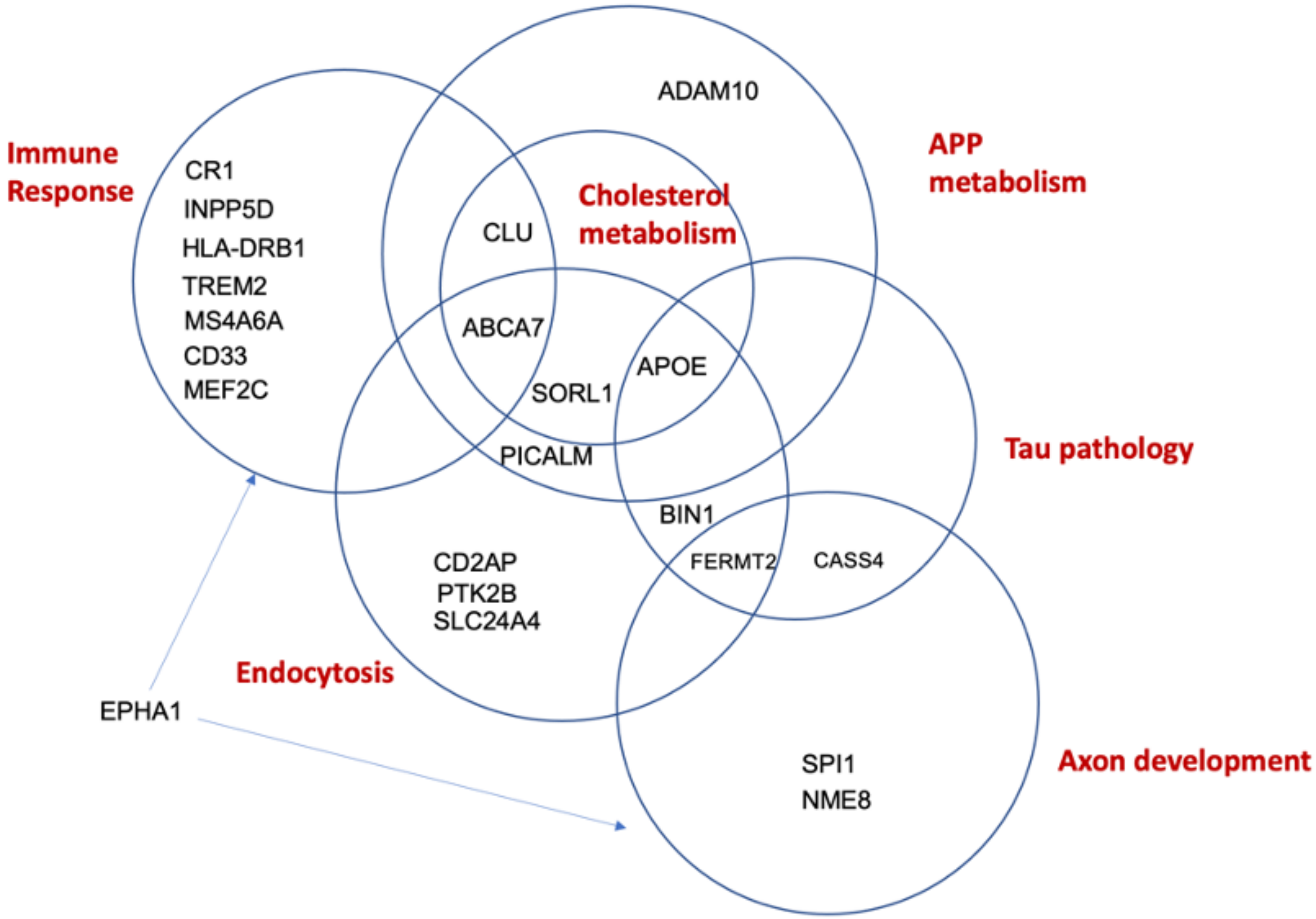


*EPHA1 is included in both the Immune response and Axon development pathways

**Supplementary Figure 2. Model predicted simple slope of p-PRSs/PRS on cognition at different age with 95% confidence interval in Wisconsin ADRC**

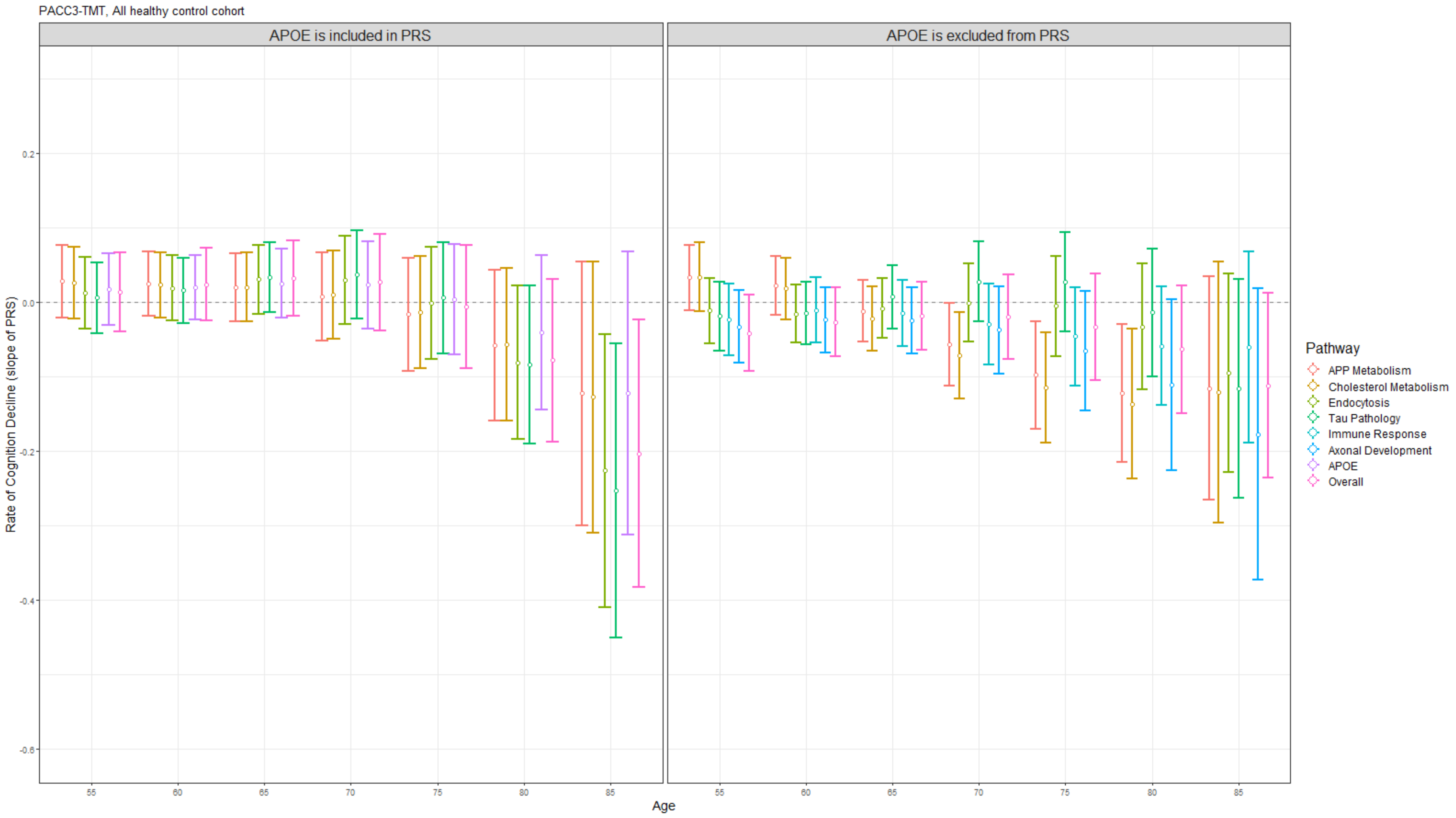


Supplementary Figure 2 presents the model predicted simple slope (based on the estimates) of p-PRSs/PRS on the PACC3-TMT score at various age points in the AHC sample extracted from the Wisconsin ADRC with a 95% confidence interval. Within each figure, the left panel depicts the simple slope estimates of p-PRSs/PRS, excluding *APOE* score, and the simple slope estimates of p-PRSs/PRS including *APOE* score are shown in the right panel. *APOE* is not theoretically affecting immune response and axonal development pathways, so the simple slope estimates of p-PRSs/PRS of these two pathways are only shown in the left panel. All association analyses are performed using the linear mixed effect model and adjusted for within-individual. In addition to p-PRSs/PRS, age (linear, quadratic, and cubic), and their interactions, additional covariates include gender, education years, and practice effect. Simple slope estimates were calculated based on parameters from the linear mixed effect model.

**Supplementary Figure 3. Model predicted simple slope of p-PRSs/PRS on biomarker at different age with 95% confience interval in Wisconsin ADRC**

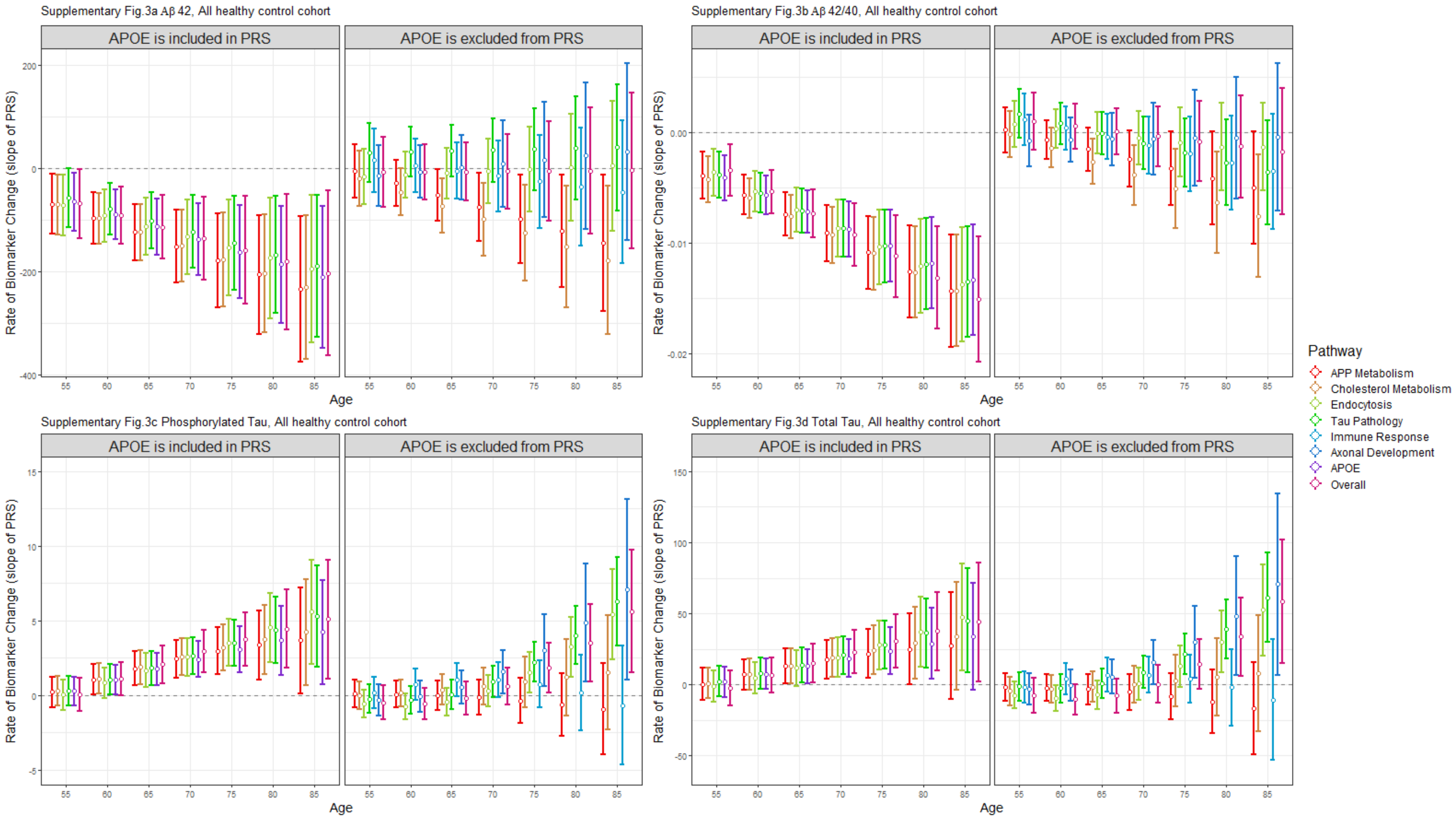


Supplementary Figure 3 presents the model predicted simple slope (based on the estimates) of p-PRSs/PRS on beta-amyloid 42 (Figure 3a), beta-amyloid 42/40 ratio (Figure 3b), phosphorylated tau (Figure 3c), and total tau (Figure 3d) at various age points in the AHC sample extracted from the Wisconsin ADRC with a 95% confidence interval. Within each figure, the left panel depicts the simple slope estimates of p-PRSs/PRS, excluding *APOE* score, and the simple slope estimates of p-PRSs/PRS including *APOE* score are shown in the right panel. *APOE* is not theoretically affecting immune response and axonal development pathways, so the simple slope estimates of p-PRSs/PRS of these two pathways are only shown in the left panel. All association analyses are performed using the linear mixed effect model and adjusted for within-individual correlation. The functional form of age for all biomarker analyses is determined by spaghetti plots. In addition to p-PRSs/PRS, age, and their interactions, additional covariates include gender, and education year. Simple slope estimates were calculated based on parameters from the linear mixed effect model.
